# Implementation and adaptation of WASH FIT in healthcare facilities: A systematic scoping review

**DOI:** 10.1101/2024.08.15.24312073

**Authors:** Sena Kpodzro, Ryan Cronk, Hannah Lineberger, Lauren Lansing, Darcy M. Anderson

## Abstract

Environmental health services (e.g., water, sanitation, hygiene, energy) are important for patient safety and strong health systems, yet services in many low- and middle-income countries are poor. To address this, the World Health Organization (WHO) and the United Nations Children’s Fund (UNICEF) developed the Water and Sanitation for Health Facility Improvement Tool (WASH FIT) to drive improvements. While widely used, there is currently no systematic documentation of how WASH FIT has been adapted in different contexts and the implications of these adaptations. We conducted a systematic scoping review to assess WASH FIT adaptation and implementation, specifically evaluating context and implementing stakeholders, the WASH FIT process and adaptation, and good practices for implementation. Our search yielded 20 studies. Implementation was typically government-led or had a high level of government engagement. Few details on healthcare facility contexts were reported. Adaptation was widespread, with nearly all studies deviating from the five-step WASH FIT cycle as designed in the WHO/UNICEF manual. Notably, many studies conducted only one facility assessment and one or no rounds of improvement. However, reporting quality across studies was poor, and some steps may have been conducted but not reported. Despite substantial deviations, WASH FIT was favorably described by all studies. Good practices for implementation included adequate resourcing, government leadership, and providing WASH FIT teams with sufficient training and autonomy to implement improvements. Low-quality reporting and a high degree of adaptation make it challenging to determine how and why WASH FIT achieves change. We hypothesize that healthcare-facility level action by WASH FIT teams to assess conditions and implement improvements has some effect. However, advocacy that uses WASH FIT indicators to highlight deficiencies and promotion of WASH FIT by WHO and UNICEF to pressure governments to act may be equally or more powerful drivers of change. More rigorous evidence to understand how and why WASH FIT works is essential to improve its performance and inform scale-up.

**Highlights:**

- Adaptation in WASH FIT programs is common but not always evidence-based
- Some programs initiate WASH FIT assessments but never implement improvements
- Some programs complete one round of improvements but never iterate the WASH FIT cycle
- Low-quality evidence impedes research to identify WASH FIT’s theory of change
- WASH FIT may drive change through national advocacy and peer pressure to unlock funding

## 1 Introduction

Environmental health services (e.g., water, sanitation, hygiene, cleaning, waste management, energy) are important for safe healthcare delivery and a priority for reducing maternal and child mortality (Velleman et al., 2014). They contribute to reducing healthcare-acquired infections and antimicrobial resistance (Watson et al., 2019; WHO et al., 2020); improving patient satisfaction and care-seeking (Bouzid et al., 2018; Fejfar et al., 2021); and improving healthcare worker satisfaction, retention, and quality and efficiency of care (Anderson et al., 2023; Fejfar et al., 2021).

To accelerate progress toward universal access to environmental health services in healthcare facilities, the World Health Organization (WHO) and United Nations Children’s Fund (UNICEF) developed the Water and Sanitation for Health Facility Improvement Tool (WASH FIT) to help plan and prioritize environmental health service improvements. WASH FIT is a risk-based, continuous quality improvement tool that aims to help healthcare facilities plan, prioritize, implement, and monitor improvements to environmental health services using a participatory approach (WHO/UNICEF, 2022f). Healthcare facilities form WASH FIT teams that conduct a five-step cycle (Table 1); cycles are designed to be iterated every six to twelve months.

WASH FIT has been implemented in 75 countries and has been adopted as an important component of national strategies for environmental health services in healthcare facilities in at least 13 (WHO/UNICEF, 2024). It is designed to apply broadly across low-resource settings but recognizes the need for tailoring across contexts. The WASH FIT manual provides guidance for adapting indicators used in healthcare facility assessment forms. However, it provides little specific guidance about adapting other aspects of the improvement cycle and overall lacks specific recommendations on what, how, and when to adapt (WHO/UNICEF, 2022f).

When programs are scaled up across different contexts, adaptation is important to ensure that interventions and implementation strategies are optimized for their implementing organizations, beneficiary populations, and contexts. Improperly executed adaptations can reduce program effectiveness (Evans et al., 2021; Movsisyan et al., 2021). Given WASH FIT’s widespread use across diverse contexts, adaptation is likely necessary and commonplace. Yet, there is currently no systematic documentation of how WASH FIT has been adapted nor the implications of these adaptations. Some limited case studies have been published (WHO/UNICEF, 2022g) but provide primarily descriptive accounts of WASH FIT adaptation without rigorous systematic evaluation or comparison.

**Table 1.** Five steps of the WASH FIT improvement cycle as described in the WHO/UNICEF WASH FIT manual (WHO/UNICEF, 2022f)

| **WASH FIT step** | **Description** |
| --- | --- |
| 1. Establish and train the WASH FIT team; document decision-making | Form a multidisciplinary team of healthcare facility staff and external stakeholders who will lead and implement WASH FIT. Assign and document roles and responsibilities for each team member. |
| 2. Assess the facility | Use the contextualized WASH FIT assessment forms to collect data on the facility's WASH services and infrastructure status. |
| 3. Identify and prioritize areas for improvement | Identify and prioritize the risks related to WASH that affect the healthcare facility's quality of care and patient safety. |
| 4. Develop an improvement plan and take action | Based on the assessment and risk analysis, develop a time-bound incremental action plan to address the gaps and improve the WASH services and infrastructure in the healthcare facility. |
| 5. Monitor, review, adapt, and improve | Implement the improvement plan and continuously monitor the progress and impact of the actions taken; share data on the healthcare facility, local, and national levels. |

We conducted a systematic scoping review to understand how WASH FIT has been implemented and adapted in different contexts. Our findings can be used to understand different contexts where WASH FIT is implemented, how it is being adapted within those contexts, and lessons learned and best practices that can be used to improve future implementation. Our specific research objectives were to (1) describe contexts and stakeholders involved in WASH FIT implementation, (2) describe the WASH FIT process and identify any adaptations, and (3) assess good practices for implementing WASH FIT.

## 2 Methods

### 2.1 Search Strategy

We conducted a systematic scoping review to identify peer-reviewed and grey literature studies describing the implementation and adaptation of WASH FIT in healthcare facilities in low- and middle-income countries. We searched two academic databases, PubMed and Scopus, on 13 February 2024 for titles, abstracts, and keywords using the following terms: “WASH FIT,” “WASH FAST,” “Water and sanitation for facility improvement tool,” and “Water and sanitation for health facility improvement tool.” We conducted all searches in English. Searches were restricted to literature published from 2017 onwards (when the first edition of WASH FIT was published).

We searched for grey literature on washinhcf.org (the largest online repository of resources related to environmental health services in healthcare facilities, curated by the WHO and UNICEF) on 6 February 2024. Resources are “tagged” with keywords by authors at the time of upload. We screened all the resources tagged with the keyword “WASH FIT.” Additionally, we scanned the references of a 2022 report on WASH FIT case studies and consulted with WHO and UNICEF experts to identify additional relevant studies.

### 2.2 Eligibility Criteria and screening

We included studies that described the implementation and/or adaptation of WASH FIT in at least one healthcare facility in a low- or middle-income country. We required that the studies provide at least some description of the healthcare facilities where WASH FIT was implemented, such as their location, size, type, or level of care. We considered studies written in English, French, or Spanish of the first or second editions of WASH FIT, regardless of their publication status or study design. We excluded studies that reported only on pre-implementation steps, such as national training of trainers but did not describe any WASH FIT implementation in a healthcare facility. We excluded studies incorporating WASH FIT indicators for research and monitoring but never implementing a WASH FIT program.

In some instances, we found reports on washinhcf.org that described the same WASH FIT program as later reports or peer-reviewed literature. In these cases, we included both only if they presented meaningfully different information. For example, where we found baseline and endline reports presenting the same data, we included only one. Where we found follow-up studies of the same program site that reported on different outcomes, we included both. We included the most recent publication in cases where we did not include both studies. A single reviewer screened all studies at the title and abstract stage. For full-text screening, each study was screened by two reviewers, and disputes were resolved by discussion.

### 2.3 Data Extraction and Synthesis

A team of five authors developed a preliminary extraction form using Microsoft Excel. Two authors pilot-tested the form with three of the included studies, compared results, and refined it based on consensus. Two authors independently extracted each study. A third author compared and reconciled the results as necessary. The final extraction form is included in Supplemental Information File 1.

We synthesized data from the included studies according to the three study objectives. To describe WASH FIT contexts and implementing stakeholders, we compiled location and facility characteristics. To describe WASH FIT processes, we extracted data on which of the five WASH FIT cycle steps a study completed, plus any additional steps not described in the official WASH FIT manual. We then compared the reported steps in each study against the process as described in the WASH FIT manual (WHO/UNICEF, 2022f) and documented any adaptations. To assess good practices for implementing WASH FIT, we first compiled self-described success factors, challenges, barriers, facilitators, or other language suggesting best practices, lessons learned, or recommendations based on implementation experience. Once compiled, we grouped similar information to distill common themes.

## 3 Results

### 3.1 Included literature

Our search retrieved 176 total articles, of which 165 were unique. After title and abstract screening, we reviewed 36 full texts. We included twenty studies in the final extraction (Figure 1).

**Figure 1.**
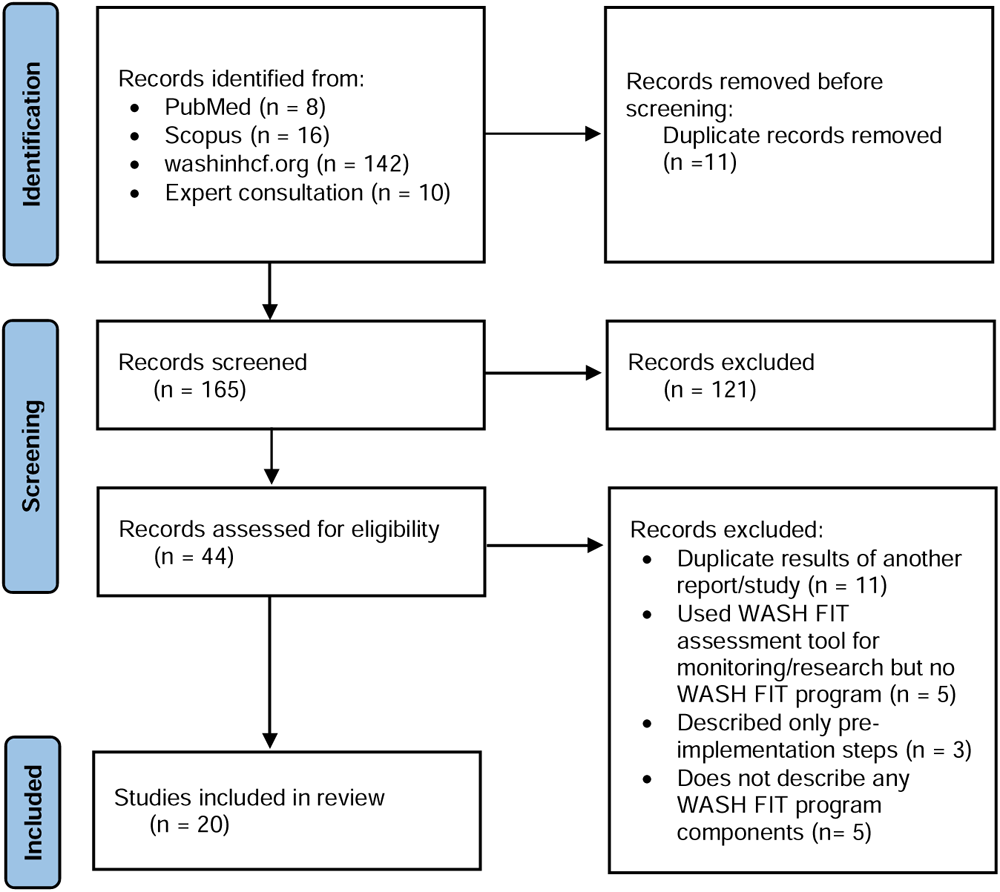
Flow chart of studies considered in this systematic review of WASH FIT adaptation.

### 3.2 Contexts and stakeholders for WASH FIT implementation

Literature from our search represented approximately a fifth of the countries known to implement WASH FIT as of 2024. Most studies were from the African and Southeast Asian regions (Figure 2).

**Figure 2.**
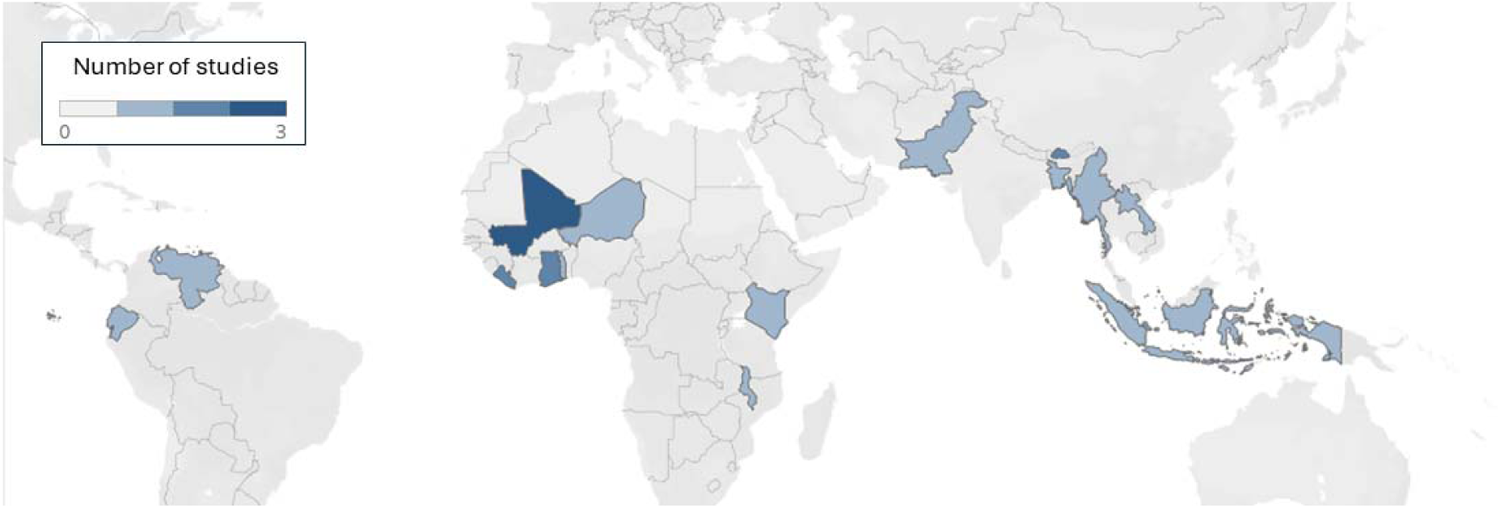
Countries with WASH FIT studies included in this systematic review of WASH FIT adaptation.

Across all studies, WASH FIT was implemented in 719 healthcare facilities, with an average of 36 facilities per study (ranging from 1 to 256). “Hospitals” and “health centers” were the most used terms to describe facilities. Most studies reported little information about the healthcare facilities where WASH FIT was implemented. Three studies specified the number of beds per facility (ranging from 4 to 650); one study provided information on the number of patients per month. Qualitative descriptors were more commonly reported but still missing in many studies (Table 3).

**Table 3.** Characteristics of healthcare facilities included in a study on WASH FIT adaptation.

| **Qualitative facility descriptors** | **Studies reporting characteristic (n, %)** | **Studies reporting no information (n, %)** |
| --- | --- | --- |
| *Location* |  | 6 (30%) |
| Rural | 5 (25%) |  |
| Urban | 4 (20%) |  |
| Emergency | 2 (10%) |  |
| Mixed urban and rural areas | 2 (10%) |  |
| *Highest level of care provided* |  | 13 (65%) |
| Primary | 3 (15%) |  |
| Secondary | 2 (10%) |  |
| Tertiary and above | 2 (10%) |  |
| *Facility management and ownership* |  | 9 (45%) |
| Public | 8 (40%) |  |
| Private | 2 (10%) |  |
| Both public and private within the study sample | 1 (5%) |  |
| *Type of patient services provided* |  | 13 (65%) |
| Inpatient | 6 (30%) |  |
| Outpatient | 5 (25%) |  |
| Maternity | 5 (25%) |  |

WASH FIT programs were mostly led by Ministries of Health or similar health-related government agencies (e.g., national public health institutes and regional health departments), with 14 studies (70%) reporting leadership or substantial involvement from the government. Of these, most (10 studies) had support from another agency, most commonly the WHO, UNICEF, or an international non-governmental organization (NGO). The remaining six studies reported that WASH FIT programs were implemented by NGOs (n=2 studies) or did not describe the implementing stakeholders (n=4 studies).

At the healthcare facility level, WASH FIT teams typically included at least healthcare workers, administrators, or healthcare facility managers, and infection prevention and control or WASH focal points where available. Some facilities included additional personnel, such as cleaners, community representatives, regional health department staff, and/or NGO staff. Nine studies did not describe the WASH FIT teams, though six indicated that teams had been formed or planned.

In three studies, WASH FIT teams were formed at the program level of medical, public health, or research professionals that conducted activities across multiple healthcare facilities in the program (Abdoulbaki Illio, 2023; Maina et al., 2019; WASH FIT supportive supervision, 2020). In these studies, there was no mention of WASH FIT teams formed at the facility level.

### 3.3 WASH FIT Process and Adaptations

We documented the WASH FIT processes reported by studies and compared them to the five-step cycle described in the WHO/UNICEF WASH FIT manual. We identified three studies that completed all five steps in accordance with the WASH FIT manual (Report on Piloting of WASH FIT, 2021; Teme, 2023; Weber et al., 2018). The remaining 17 modified either the number, order, and/or activities of steps in some way.

Eleven studies performed at least three WASH FIT steps (Ashinyo et al., 2021; Aung & Swinchatt, 2019; Dorji, 2020; Kabir et al., 2023; Ogando dos Santos et al., 2021; Person et al., 2020; Sehar et al., 2022, 2022; WHO/UNICEF, 2022d, 2022b, 2022c). Five studies performed no more than two WASH FIT steps (Abdoulbaki Illio, 2023; Doku et al., 2022; Maina et al., 2019; WASH FIT supportive supervision, 2020; WHO/UNICEF, 2022a). Two studies reported no information about the steps taken (Kanagasabai et al., 2021; Nyirenda & Ferrey, 2018).

Every study conducted Step 2 (facility assessment). The least common step was Step 5 (monitor, review, adapt) (Figure 3). It was difficult to determine how many times studies iterated the WASH FIT cycle, if at all. Ten studies stated the number of assessment cycles (ranging from 1 to 3, with 1 being the most common). Three studies did one round of improvement; four did none. Thirteen studies did not state the precise number of improvement rounds, though some suggested that the WASH FIT cycle had been iterated more than once.

**Figure 3:**
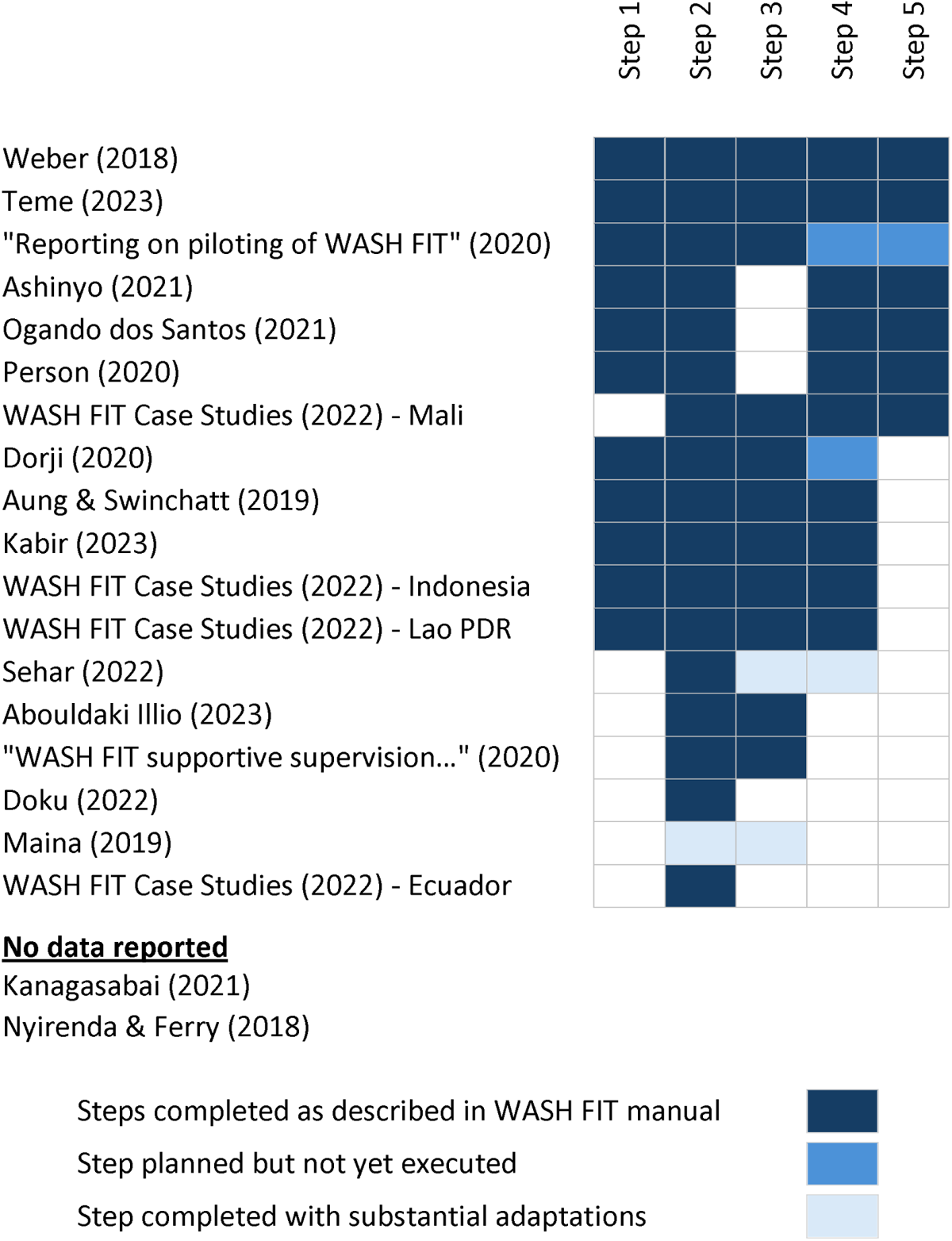
WASH FIT steps reported by each included study.

By comparing the similarities and differences in the steps, we identified two adaptation archetypes: “WASH FIT assessors” and “WASH FIT improvers.” WASH FIT assessors conducted Step 2 (facility assessment) and sometimes Step 3 (risk-based improvement planning) but did not report forming WASH FIT teams or implementing improvements. WASH FIT improvers conducted Steps 2 and 4 (implement improvements) but did not report conducting at least one step—most commonly Step 5 (monitor, review, adapt) or Step 3. Figure 4 depicts the adapted WASH FIT process for each archetype.

**Figure 4.**
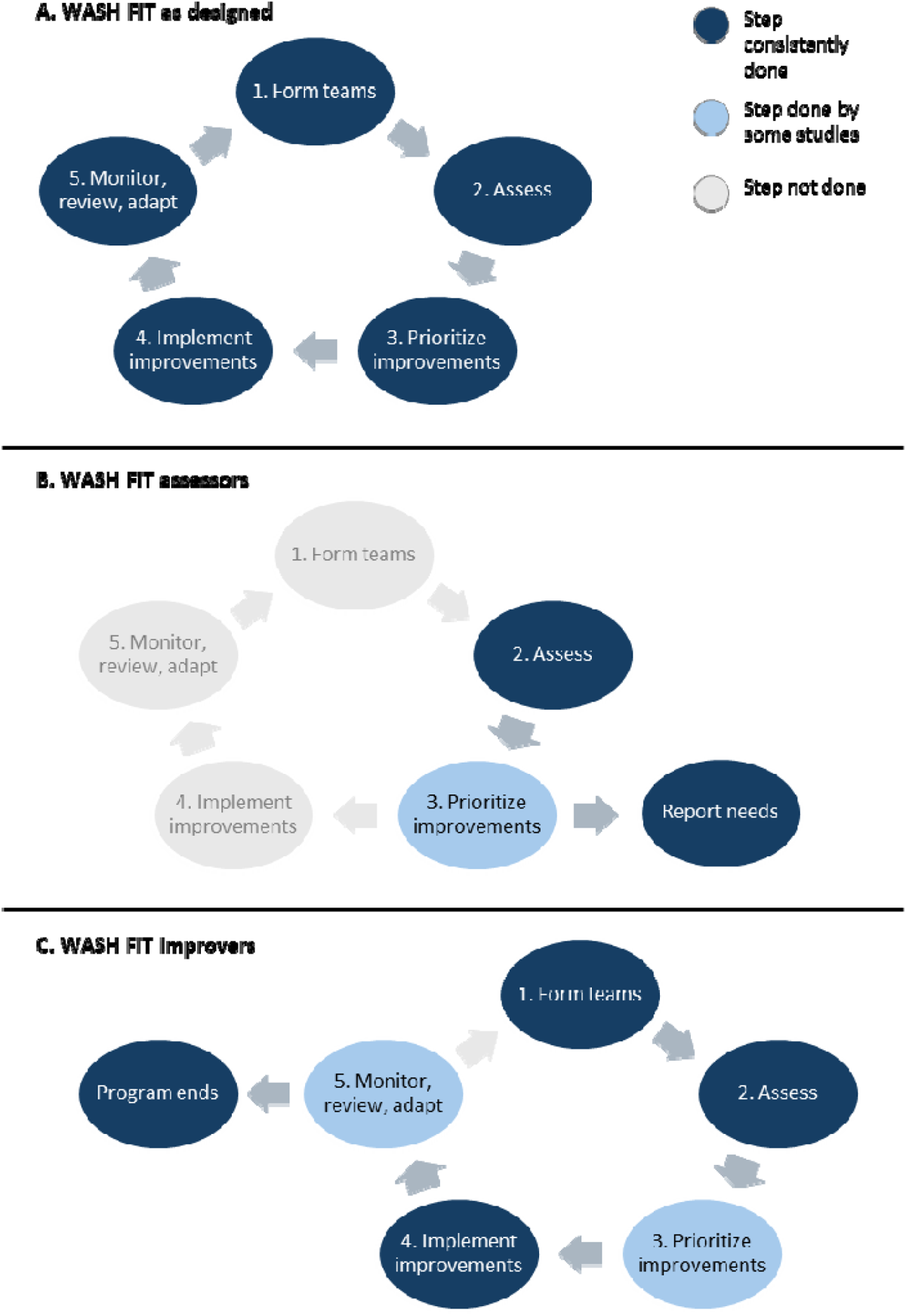
WASH FIT archetypes. Panel A: WASH FIT as designed (performing all five steps); Panel B: WASH FIT initiators (performing Steps 2 and 3); Panel C: WASH FIT improvers (performing Step 4 and at least two other steps).

#### 3.3.1 WASH FIT assessors

We identified five studies that we categorized as WASH FIT assessors (Abdoulbaki Illio, 2023; Doku et al., 2022; Maina et al., 2019; WASH FIT supportive supervision, 2020; WHO/UNICEF, 2022a). These studies assessed the facility and sometimes created a risk-based improvement plan based on the assessment. However, no further effort was reported to execute improvements once these steps were taken.

Sometimes, the results of the assessment were used to demonstrate the need for improvements to external stakeholders (e.g., donors and government agencies) and advocate for resources and investments. For example, Doku et al. (2022) conducted a WASH FIT assessment and concluded that the assessment revealed a need for better WASH programming. The study described aspirations to establish a WASH FIT team to conduct quarterly assessments to monitor needs and activities but did not necessarily plan to follow other aspects of WASH FIT to prioritize and execute specific improvements based on the assessment results.

In other cases, WASH FIT assessments highlighted improvements already made. For example, a WHO/UNICEF case study from Ecuador reported that WASH FIT assessments were done to “draw attention and understand the status of WASH in healthcare facilities” and to raise awareness of improvements in water supply and treatment that were done previously as part of a COVID-19 response program (WHO/UNICEF, 2022a).

While all studies that we classified as WASH FIT assessors reported that they had conducted WASH FIT or established a WASH FIT program, no studies in this archetype reported forming a WASH FIT team at the healthcare facility with training and autonomy. Often, what was described instead was activities to form or build the capacity of regional teams. For example, Illio (2023) describes that a “WASH/IPC team” was formed at the district level, who conducted one visit to each healthcare facility to carry out the WASH FIT assessment. Another program in Liberia established a “supervision and mentorship” program. WASH FIT assessments were done by a program-level team as part of establishing the WASH FIT program. Individual healthcare workers were paired into a “supervision and mentorship” program, and the study recommended—but did not establish—facility-level WASH FIT teams (WASH FIT supportive supervision, 2020).

#### 3.3.2 WASH FIT improvers

We identified ten studies that we categorized as WASH FIT improvers (Ashinyo et al., 2021; Aung & Swinchatt, 2019; Dorji, 2020; Kabir et al., 2023; Ogando dos Santos et al., 2021; Person et al., 2020; Sehar et al., 2022; WHO/UNICEF, 2022d, 2022b, 2022c). These studies performed Step 4 (implement improvements) and at least two other steps of the WASH FIT cycle but with some deviation reported from the WASH FIT manual.

The most common adaptation among WASH FIT improvers was that studies did not report conducting Step 5 (monitor, review, adapt). In some cases, WASH FIT was done to respond to a particular acute need (e.g., COVID-19 response) and not intended for long-term activities (Ashinyo et al., 2021). In other cases, studies were a six-month or one-year progress report that contained a pre-implementation baseline and six-month or one-year evaluation of the WASH FIT assessment indicators, with at least one improvement cycle (Aung & Swinchatt, 2019; Dorji, 2020). Whether programs continued activities beyond the one-year assessment point is not reported but plausible. Two studies reported quasi-experiment designs with a baseline and endline evaluation that focused on changes in WASH FIT assessment indicators over a fixed period at the end of the project grant cycle (Kabir et al., 2023; Person et al., 2020). In these cases, studies implied that engagement from external research teams or technical support agencies ended after the endline. However, activities from healthcare facility-level WASH FIT teams may have continued beyond official program support but were not reported.

The second most common adaptation was that studies did not report conducting Step 3 (risk assessment). All studies in this archetype conducted Step 2 (facility assessment). The WASH FIT manual recommends that, after the assessment, healthcare facilities develop a tailored improvement plan that prioritizes improvements based on need (i.e., low-scoring indicators) but also maximizing impacts on health and wellbeing, feasibility to correct, and other priorities. However, some studies reported moving directly from the assessment into implementing improvements without a dedicated risk assessment step. For these studies, we were not able to determine what criteria were used to select specific improvements (Ashinyo et al., 2021; Ogando dos Santos et al., 2021; Person et al., 2020). In one case of a particularly extensive adaptation, WASH FIT was used to assess baseline conditions and justify the need for intervention. The study team then designed an intervention package based on the most common needs across all study facilities, which they uniformly delivered across the study area, rather than tailoring improvement approaches based on facility-specific risk assessments. Iterative WASH FIT assessments were conducted throughout the study period as performance indicators but did not explicitly inform intervention activities after the baseline assessment (Sehar et al., 2022).

### 3.4 Reported best practices for implementation

We compiled self-reported good practices for WASH FIT implementation and compared them across studies to distill common themes. This analysis identified four thematic areas supporting successful implementation, as described below.

#### 3.4.1 Adequate resourcing

The most commonly described success factor for implementing WASH FIT was adequate financial resources (Ashinyo et al., 2021; Doku et al., 2022; Dorji, 2020; Kabir et al., 2023; Maina et al., 2019; Nyirenda & Ferrey, 2018; Report on Piloting of WASH FIT, 2021; Teme, 2023; WASH FIT supportive supervision, 2020; Weber et al., 2018). Steps 1-3 to form WASH FIT teams, assess facilities, and develop improvement plans based on risk assessments were described as requiring minimal cost or resource inputs. However, implementing improvement plans required substantial investment or in-kind support (e.g., constructing infrastructure and providing supplies). Several studies reported that facilities had developed improvement plans that they could not implement because there was no dedicated funding for improvements (Kabir et al., 2023; Nyirenda & Ferrey, 2018; Weber et al., 2018). In contrast, WASH FIT programs that included funding for facility-level improvements faced fewer barriers to implementing improvement plans (Aung & Swinchatt, 2019; Person et al., 2020).

#### 3.4.2 Government leadership and coordination

Multiple studies reported the benefits of leadership and coordination with the government and the Ministry of Health (Dorji, 2020; Kabir et al., 2023; Maina et al., 2019; Report on Piloting of WASH FIT, 2021; Teme, 2023; Weber et al., 2018). National-level government engagement was important to show support for WASH FIT and create motivation and accountability across different levels of WASH FIT programs (Weber et al., 2018). Local government support was important for the accountability and motivation of healthcare facility staff, and programs with little interaction and support from local governments struggled to mobilize healthcare facility teams to uptake and sustain WASH FIT activities (Dorji, 2020; Kabir et al., 2023; Teme, 2023). Engagement with local governments also helped ensure that action plans were realistic and adequately resourced, particularly when programs did not receive external non-governmental support for improvements (Report on Piloting of WASH FIT, 2021).

#### 3.4.3 Training, coordination, and autonomy of facility-level WASH FIT teams

Successful WASH FIT teams had strong training and assigned roles and responsibilities, which were ideally coordinated with their other duties (Doku et al., 2022; Nyirenda & Ferrey, 2018; Teme, 2023). WASH FIT team members sometimes sat on multiple committees, which caused confusion and competing or duplicative effort when not properly coordinated (Nyirenda & Ferrey, 2018). WASH FIT was described as time-consuming, and without clear responsibility or belief in its importance, staff sometimes reported low commitment (Doku et al., 2022; Ogando dos Santos et al., 2021; Weber et al., 2018). Multiple studies mentioned challenges related to the autonomy or decision-making power of WASH FIT teams, where individuals on the WASH FIT team were not authorized to take action or delegate activities needed to carry out improvement plans (Kabir et al., 2023; Teme, 2023), or where a strong hierarchy within the healthcare facility hindered action (Weber et al., 2018). One study suggested that WASH FIT teams may be aided by creating standard operating procedures for operations and maintenance to anticipate and authorize common tasks (Kabir et al., 2023).

#### 3.4.4 Mentoring and feedback for facility-level teams

Supportive supervision and onsite mentorship improved WASH FIT implementation (Teme, 2023; WASH FIT supportive supervision, 2020). One program established a supplementary peer mentoring program to strengthen the knowledge and capacity of healthcare workers related to environmental health, which was not a dedicated part of the WASH FIT cycle but strengthened activities (WASH FIT supportive supervision, 2020). Other studies mentioned opportunities for self-evaluation to strengthen the capacity of WASH FIT teams (Doku et al., 2022). Strong supervision of WASH FIT teams, both within the healthcare facility and from program-level staff, was reported to improve performance (Doku et al., 2022; Person et al., 2020; Teme, 2023; Weber et al., 2018).

## 4 Discussion

We conducted a systematic scoping review to assess implementation and adaptation of WASH FIT. Overall, implementation was typically government-led or had a high level of government engagement. Few details on healthcare facility contexts were reported. Adaptation was widespread, with nearly all studies deviating from the five-step WASH FIT cycle as designed in the WHO/UNICEF manual (WHO/UNICEF, 2022f). Notably, many studies conducted facility assessments and one or no rounds of improvement. However, reporting quality across the literature was poor, and some steps may have been conducted but simply not reported. Despite substantial deviations, WASH FIT was favorably described by all studies.

Low quality reporting and a high degree of adaptation makes it difficult to determine how and why WASH FIT works to achieve change. However, we hypothesize that the theory of change as proposed in the WHO/UNICEF manual is likely incomplete—WASH FIT likely works in ways that are not well understood. Existing literature offers some clues, but there is a considerable gap in rigorous, well-documented evidence to fully identify and evaluate a theory of change. We propose several hypotheses below.

Understanding how and why WASH FIT achieves impact is critical to ensuring it is implemented effectively. Without this, country governments and organizations miss opportunities to learn and improve, implement in contexts with the greatest chance of success, and tailor to other contexts as necessary and appropriate. We point to cautionary evidence from household water and sanitation programs, in which community-led total sanitation was rapidly scaled up across dozens of countries because it was perceived as fast, low-cost, and effective. However, this scale-up was based on early evidence from a very limited range of contexts. Subsequent research suggested that such wide-spread scale-up may have been inappropriate and that the approach is suited for successful implementation in a narrower range of contexts (Israel et al., 2022; Stuart et al., 2021; Zuin et al., 2019). These learnings came only after billions of dollars of investment and a series of trials demonstrating poor performance (Barnard et al., 2013; Cameron et al., 2019; Clasen et al., 2014; Patil et al., 2014).

WASH FIT has followed a similar trajectory of rapid scale-up. First released in 2017, as of 2024, it is now implemented in over 75 countries, with billions of dollars of investment. WASH FIT is well-liked and perceived as a low-cost, universal solution. Yet this review and a 2024 systematic review on WASH FIT effectiveness (Lineberger et al., unpublished manuscript) are the first evidence synthesis efforts—and both reviews are hindered by the poor quality of included studies. Our assessment of good practices suggests that WASH FIT is not universally suited to all contexts and performs better in contexts with several key factors (e.g., adequate resourcing and local government engagement).

Given that further scale-up seems likely, we call for more rigorous evidence to answer how and why WASH FIT works and in what contexts. The final section of our discussion suggests specific strategies for strengthening future studies.

### 4.1 Continuous quality improvement principles underpin WASH FIT’s theory of change

The WHO/UNICEF WASH FIT manual does not specifically articulate a theory of change to explain how or why WASH FIT programs achieve impact, though one study has proposed a logic model of inputs, outputs, outcomes, and impacts (Weber et al., 2019). The manual does state that WASH FIT evolved out of water safety plans, which are a risk-based approach to safeguarding and improving drinking water quality promoted by the WHO for household drinking water (WHO, 2023). WASH FIT extends the approach to include additional environmental health services and tailors the process to the context and stakeholders of a healthcare facility.

Though not branded as such, at its core, WASH FIT (and its predecessor, water safety plans) are continuous quality improvement (CQI) approaches. CQI evolved in the manufacturing sector to improve production lines. It has since been widely applied in health and other sectors (Knudsen et al., 2019; Nicolay et al., 2012; Taylor et al., 2014; Zamboni et al., 2020). Its core functions are engaged teams committed to ongoing, iterative efforts to identify and execute improvements based on measurable performance metrics. The intent is that improvements are rapid and incremental, under the theory that smaller and faster changes are easier to execute, evaluate, and course-correct as needed (Knudsen et al., 2019; Taylor et al., 2014). Specific improvements are not dictated as part of the overall CQI approach but rather designed or tailored to the specific context by the project team based on specific performance indicators and needs assessments.

CQI approaches are popular in the healthcare sector, though rigorous evidence underpinning their effectiveness is mixed. While CQI has been applied in the healthcare sector for decades, systematic reviews from the past five years still deem the evidence of its effectiveness “uncertain” (Hill et al., 2020), with other reviews suggesting that CQI may only be appropriate for specific contexts (Taylor et al., 2014; Walshe & Freeman, 2002). Application of CQI to address environmental health services challenges is uncommon and primarily based in community settings, though existing studies have demonstrated positive preliminary results (Anderson et al., 2021; Fisher et al., 2020).

Adherence to CQI principles likely mediates effectiveness, with studies finding that metrics like meeting frequency and level of commitment from the CQI team are critical success factors (Endalamaw et al., 2024; Hill et al., 2020). One criticism is that programs commonly adopt the “CQI” label but are not true CQI programs, because they fail to complete more than one improvement cycle or carry assessment data forward to inform action—thereby violating core CQI functions (Taylor et al., 2014).

### 4.2 WASH FIT adaptation suggests low implementation fidelity and gaps in the theory of change

Our results suggest that—while some programs implement WASH FIT as intended in the original WHO/UNICEF manual—there is considerable adaptation. Some of this adaptation is intentional, such as refining assessment indicators to suit larger, multi-ward facilities, using a well-documented, evidence-based process (Maina et al., 2019). However, we believe that much of the adaptation is better characterized as programmatic “drift” (i.e., deviation from the original program design that is not intentionally planned following evidence). Much of the adaptation we observed involved initiating but not completing the full WASH FIT cycle or completing the cycle only once without iterative improvement. These adaptations contradict the CQI principles on which WASH FIT is based.

Given the widespread adaptation of WASH FIT that often contradicts core functions of CQI, we pose two questions: Are adapted WASH FIT programs effective, and if so, why? What theory of change underpins their impact pathways? For the first question on effectiveness, a 2024 systematic review on WASH FIT effectiveness—which included these adapted programs—found that improvements to WASH services (e.g., infrastructure access, functionality) were plausible, though study designs were weak and the overall quality of evidence was poor. Evidence from other sectors indicates that adaptations that deviate from the evidence-based core functions of an intervention often undermine effectiveness (Evans et al., 2021; Movsisyan et al., 2021). Yet despite violating supposed core principles based on CQI, WASH FIT was favorably viewed in every article we included in this study, with nearly all making recommendations for continuing and expanding WASH FIT programs.

This brings us to the second question: if WASH FIT is perceived as effective and preliminary evidence does not refute that perception, why is it effective? How does it achieve change? For WASH FIT teams that adhere to CQI principles, this may drive changes in environmental health services at the facility level. However—for the many programs that do not—we hypothesize that CQI principles may not be the primary driver of change or, at least, are complementary to multiple impact pathways that work in different combinations across different contexts.

One possible change pathway is through monitoring and advocacy. WASH FIT provides the most comprehensive, structured tool to document WASH conditions and a straightforward way to rank and communicate risks and needs to improve. The WHO and UNICEF have developed extensive training, handbooks, and other supporting documents in multiple languages. WASH FIT represents one of the most comprehensive and user-friendly assessment tools, offering substantial advantages over other tools. Few comprehensive assessment tools exist, and those that do are primarily larger national health monitoring surveys with considerably fewer environmental indicators and little to no training materials or user support (e.g., (USAID, 2018; WHO, 2019; WHO/UNICEF, 2018)). We hypothesize that this makes WASH FIT an appealing entry point for countries and organizations that are planning their programs with a situation assessment. In support of this hypothesis, we point to studies that cited WASH FIT assessment results as the key factor in national-level advocacy efforts that unlocked substantial funding (WHO/UNICEF, 2022g). With rigorous tools and protocols to document and communicate needs, stakeholders can more effectively advocate for resources to address them. In this case, the primary change mechanism is the data-backed advocacy activities that unlock investments, which can be deployed to improve conditions with or without WASH FIT teams acting at the facility level.

Another possible change pathway is through peer pressure created by successful branding and promotion of WASH FIT by key stakeholders. The WHO and UNICEF’s leadership in developing and promoting WASH FIT lends credibility and legitimacy. The WHO/UNICEF 2024 Global Framework for Action on WASH, Waste and Electricity in Healthcare Facilities lays out actions to achieve the 2030 development agenda, with promoting and scaling-up WASH FIT as a key priority (WHO/UNICEF, 2024). Accompanying this document is a consensus statement, in which country governments, multilateral agencies, NGOs, and other partners endorse the framework and make specific commitments to support its implementation. In this case, WASH FIT likely drives facility-level change because it pressures funders and implementers to address environmental health issues in healthcare facilities. Resources are then funneled through WASH FIT programs because of their promotion by strategic actors like WHO and UNICEF.

We suspect that several of these pathways work together to create change. The theory of change implied in the WASH FIT manual (i.e., iterative improvement based on CQI principles) may not be the primary mechanism of action. Alternative pathways to change through advocacy and peer pressure to mobilize resources may be equally or more influential.

### 4.3 Limitations of the evidence and next steps for strengthening rigor

#### 4.3.1 Develop and collect indicators on the WASH FIT process and healthcare facility context

A key limitation of this review is the reporting quality of underlying studies. For most studies that did not report completing specific steps of the WASH FIT cycle, we could not determine whether the step was truly not done or simply not reported in the study. To understand WASH FIT’s theory of change, a fundamental first step is strengthening documentation of the WASH FIT process. At a minimum, studies should indicate which of the five steps of the WASH FIT process were done at the healthcare facility-level (explicitly stating if steps were not done) and the number of assessment and improvement rounds. For studies focused on facility-level outcomes, we suggest more in-depth reporting of the facility-level WASH FIT process. Reporting guidelines are used to standardize and strengthen research reporting, assisting readers with contextualizing and interpreting the findings and enabling more comprehensive evidence synthesis through systematic reviews. Reporting guidelines for CQI studies are applicable to WASH FIT (Ogrinc et al., 2008); we suggest their use. In some instances, information on the specific improvements made at the facility level may also be pertinent. In this case, we recommend reporting guidelines for water, sanitation, and hygiene interventions (Crocker et al., 2024).

Weak reporting of the healthcare facility context is also a challenge. For example, over half of the studies in our review provided no information on the level of care or type of medical services provided by the healthcare facilities. Addressing this challenge will require definitions and indicators to describe and measure facility types, size, and other characteristics. The 2022 WHO and UNICEF report on global monitoring data for environmental health services disaggregates by “hospitals” versus “non-hospitals” (WHO/UNICEF, 2022e). This classification is based on whether a facility is self-identified as a “hospital.” The report suggests that more nuanced classifications would be beneficial. Some work has already been done within the energy sector. A 2015 review to classify healthcare facilities to develop benchmarks for energy efficiency in high-income countries defined six categories: patient type, care provided, management and ownership, level of care, facility size, and location (Ahmed et al., 2015). However, additional research is needed to identify and validate specific indicators. In the interim, we advise that WASH FIT studies provide descriptions of these six categories using quantitative indicators whenever possible.

We recommend that the WHO and UNICEF provide templates to help teams document and report the facility context and WASH FIT process as part of the assessment tools and manual. We propose that this would have several benefits. First, it would streamline data collection for facility-level teams, who would not need to manage separate assessment tools and data management processes. Second, it would increase the collection of this information by including it as a default module, where teams would need to consciously opt out. Third, it would facilitate comparisons across facilities within a program and across programs by creating standard indicators like those that already exist in the facility assessment tool. Additional research will be needed to develop and pilot indicators.

#### 4.3.2 Encourage and support WASH FIT teams to conduct process-focused research

Program evaluations are often focused on impact studies. However, process-focused research is critical to understanding how and why impacts may be achieved. WASH FIT is naturally inclined to generate substantial amounts of quantitative data through repeated facility assessments, and there are tools to support quantitative data management (e.g., there is a KoboToolbox module to digitally collect WASH FIT indicators, which can interface with data visualization dashboards (WASH FIT Assessment - Kobo Toolbox Version, n.d.). WASH FIT also generates considerable qualitative information (e.g., risk assessments, improvement plans, and worksheets documenting the WASH FIT team composition and decision-making). However, these items are less likely to be systematically documented and reported. We encourage WASH FIT programs to support facility-level WASH FIT teams to collect and report these items. Designing program monitoring and data management systems to incorporate qualitative information would be a good first step. In the long-term, greater investment in monitoring and learning to analyze this qualitative process information will be needed.

## Data Availability

All data produced in the present work are contained in the manuscript

## Declarations

### Authorship contribution statement

**Sena Kpodzro**: Writing – original draft, Visualization, Analysis. **Ryan Cronk**: Writing – review and editing, Supervision, Conceptualization. **Hannah Lineberger**: Analysis, Writing - review and editing. **Lauren Lansing**: Analysis. **Darcy Anderson**: Writing – original draft, Writing – review and editing, Supervision, Conceptualization.

### Conflict of Interest

The authors declare no conflicts of interest.

### Funding

This work was funded in part by a grant from the Wallace Genetic Foundation to the Water Institute at UNC. Darcy Anderson is supported by a grant from the National Institute of Environmental Health Sciences (NIEHS) (T32ES007018).

## Notes

### Competing Interest Statement

The authors have declared no competing interest.

